# Circulating Biomarkers as Predictors of Improvement in Physical Function in Hospitalized Older Adults with Geriatric Syndromes: Findings from the REHAB-HF Trial

**DOI:** 10.1101/2024.09.13.24313662

**Authors:** Abdulla A. Damluji, Scott A. Bruce, Gordon Reeves, Amy M. Pastva, Alain G. Bertoni, Robert J. Mentz, David J. Whellan, Dalane W. Kitzman, Christopher R. deFilippi

**Author notes:** **CORRESPONDING AUTHOR**: **Christopher R. deFilippi, MD,** Vice Chair of Academic Affairs, Inova Schar Heart and Vascular, 3300 Gallows Road, Falls Church, VA 22042.

## Abstract

**Introduction:** Circulating biomarkers play an important role in patients with heart failure (HF) for risk stratification and mechanistic insights. We aimed to examine if a diverse set of biomarkers in the REHAB-HF trial would predict improvement in physical function following a 12-week tailored physical therapy rehabilitation intervention compared to attention control.

**Methods:** The study population consisted of participants ≥60 years of age who were hospitalized with acute HF and randomized to a subsequent multidomain outpatient physical rehabilitation intervention vs. attention control with outcomes of 12-week functional change including the Short Physical Performance Battery (SPPB) and six-minute walk distance (6MWD). Blood was collected prior to randomization and at 12-weeks for cardiac, renal, and inflammatory biomarkers. Linear trends across progressively higher biomarker values versus improvement in functional outcomes based on treatment assignment were evaluated. Classification and regression trees (CART) were created to estimate optimal biomarker levels associated with differential improvement in the two functional outcomes.

**Results:** A total of 242 of 349 participants (69%) had baseline biomarkers measured. In an adjusted regression model, higher baseline cardiac troponin (cTn) I and T were associated with greater gains in SPPB and 6MWD respectively with the rehabilitation intervention (*P*=0.04 and 0.03 for interaction) versus attention control. In the CART analysis of the physical rehabilitation and attention control participants, those with baseline C-reactive protein (CRP) ≥9.9 mg/L and hs-cTnT ≥36 ng/L receiving the rehabilitation intervention had a 129 m (95% CI 78-180m) greater 12-week 6MWD increase vs attention control. In contrast, for participants with CRP<9.9 mg/L there was no significant incremental 6MWD difference (30m, 95% CI -0.5m, 60.2m). For SPPB, a CRP ≥9.9 mg/L and creatinine ≥1.4 mg/dL optimally identified a differential improvement with the rehabilitation intervention versus attention control. The biomarkers (except for creatinine) decreased by 12 weeks post hospitalization but with no differences based on treatment assignment.

**Conclusion:** Higher baseline levels of biomarkers of inflammation, cardiac injury, and renal dysfunction identified older adults after a HF hospitalization with the greatest differential improvement in physical function with a rehabilitation intervention. Biomarkers may help clinicians predict the benefits of this treatment.

(Funded by the National Institutes of Health and others; REHAB-HF ClinicalTrials.gov number, NCT02196038).

## INTRODUCTION

Precision phenotyping is the process of categorizing patients based on clusters of precise, measurable, and reliable biologic or physiologic characteristics.^1^ Precision medicine in patients with cardiovascular disease (CVD) may be utilized as a method to optimize the impact of therapeutic interventions, improve health outcomes including health-related quality of life, physical function, and independence, and facilitate early intervention.^2^ The predominant treatment focus for heart failure (HF) has been on pharmacotherapies to alter the neural hormonal axis, or device based therapies to reduce the risk of sudden cardiac death or rehospitalizations.^3^ Recent data from the Rehabilitation Therapy in Older Acute Heart Failure Patients (REHAB-HF) trial found that an early, transitional, tailored, progressive multidomain physical rehabilitation intervention was associated with improved physical function and quality of life among older patients hospitalized with acute HF.^4^

A diverse set of circulating biomarkers measured in the REHAB-HF trial could potentially provide both prediction of response and mechanistic insights into how the tailored physical rehabilitation intervention can positively impact physical function after an acute HF hospitalization. In this study, we aimed to determine whether clinically available circulating biomarkers representing cardiac pathophysiology, generalized inflammation, and renal function measured pre-intervention are effect measure modifiers (i.e. identify subgroups that respond differently) to a 12-week physical rehabilitation intervention versus attention control for improvement in measures of physical function among frail older adults after an acute HF hospitalization. We also sought to examine if a rehabilitation intervention would selectively modify any of these circulating biomarker levels versus attention control as these patients recovered from an acute HF hospitalization.

## METHODS

### Study Population, Functional and Clinical Outcomes

Details of the REHAB-HF trial design, intervention, and study populations have been previously published.^4, 5^ Briefly, the study population consisted of 349 older adults ≥60 years of age who were hospitalized with acute HF and exhibited at least one symptom and two signs of HF that resulted in a change to medical therapy targeting HF domains. While there was no exclusion criteria based on left ventricular ejection fraction (LVEF), participants had to have the ability to: (1) perform basic activities of daily living prior to their index HF admission; (2) walk independently with or without assistive device for at least 4 meters prior to enrollment; (4) being discharged to home. Patients were excluded if they had: (1) end stage HF; (2) severe valvular heart disease; (3) advanced renal dysfunction; (4) dementia or severe cognitive dysfunction; (5) Planned discharge other than to home or a facility where the participant will live independently.^4^ Participants were randomized to a 12-week, three times per week tailored multi-domain rehabilitation intervention versus attention control. The primary outcome of the study was the change from the time of randomization (prior to hospital discharge) to 12-weeks in the Short Physical Performance Battery (SPPB). For this analysis we also used the change in the 6-minute walk distance (6MWD) in meters as a co-primary endpoint with its greater range of measurement potentially reflecting more granular associations with the continuous biomarker measures as well as representing different physiologic mechanisms from the SPPB.^6^ Details regarding the measurement of SPPB and 6MWD in REHAB-HF have been previously reported.^5^ Secondary outcomes of all-cause hospitalization and death were assessed at 6-months as reported in the primary study.^4^

### Biomarker Measurement

Circulating biomarkers from blood were measured at baseline prior to randomization and 12-week follow-up from serum stored at a temperature of -80°C and later thawed for the first time and analyzed simultaneously for both time points with a high sensitivity assay for cardiac troponin I (hs-cTnI, Siemens ExL, Terrytown NY) and T (hs-cTnT, Cobas e602, Roche Diagnostics, Indianapolis IN), N-terminal pro-brain natriuretic peptide (NT-proBNP, Cobas e602, Roche Diagnostics, Indianapolis IN), creatinine (Siemens ExL, Terrytown NY), and high-sensitivity C-reactive protein (hs-CRP, Siemens ExL, Terrytown NY) at the Inova Biocore laboratory (Fall Church, VA). The sex neutral 99^th^ percentile for this hs-cTnI assay is 58 ng/L and for hs-cTnT is 19 ng/L.^7^

### Statistical Analysis

Statistical analyses were performed using R statistical software version 4.3.1.^8^ For the biomarker analysis, several steps were taken to prepare the data for analysis. First, visualizations of variable distributions were developed to identify any outliers or biologically implausible values, as well as deviations from a Gaussian distribution, which are frequently observed for many of the biomarkers examined. Second, for biomarker concentrations below the limit of assay detection [LOD] (i.e. hs-cTnT, < 6 ng/L), values were imputed halfway between 0 and the LOD (i.e. 3.0 ng/L for hs-cTnT), and for biomarker concentrations above the upper limit of detection [ULD] (i.e. hs-CRP > 90 mg/L) were imputed at the ULD. Third, if non-Gaussianity was present, then a log base 2 transformation was applied so the resultant transformed variables more closely resemble a Gaussian distribution. Fourth, outlying biomarker values were excluded from subsequent analyses if they were more than 3 standard deviations away from the mean value after log base 2 transformation.

In the descriptive analysis of baseline and follow-up characteristics, categorical variables were reported as frequency (percentages) and continuous variables by mean (standard deviation), or median (25^th^, 75^th^ percentile). The frequency of missing data for each variable of interest (including baseline and follow-up biomarker measurements) was reported, and characteristics compared between those with complete vs. missing measurements of biomarkers and adjustment variables. For comparisons across treatment groups or subgroups, Wilcoxon rank sum tests and Student’s t-tests were used to compare non-Gaussian continuous variables and appropriately transformed continuous variables, respectively. Paired t-tests were used to assess differences from baseline to follow-up given the repeated measurements for each participant. Fisher’s exact tests and chi-squared tests were used to compare categorical variables. The Spearman correlation coefficients (rho) were determined for all the baseline biomarker combinations. To examine the change graphically and qualitatively from baseline to 12-weeks in each biomarker among treatment groups, the distribution of levels of each biomarker at each time point and for randomized treatment groups were displayed graphically with violin plots.

We first used a complete case analysis approach inclusive of participants with baseline biomarker measurements. Generalized linear models (GLMs) for each protein or biomarker were initially used to evaluate effect modification (multiplicative interaction) independently while controlling for demographic covariates (i.e., age, sex, race) and HF subtype (i.e., HF with preserved ejection fraction [HFpEF] defined as an EF≥45% and HF with reduced EF [HFrEF] defined as an EF<45%) that may influence treatment response. Controlling for known prognostic covariates in randomized trials has been shown to increase power and protect against chance imbalance.^9^ For continuous numeric outcomes (i.e., 6MWD), a standard linear model was used, and for binary outcomes (i.e. first rehospitalization or death by 6-months), a logistic regression model was used with outlier biomarker values removed as outlined above for both functional and clinical outcomes. Residual diagnostics were performed to investigate model adequacy, fit, and validity of model’s assumptions including normality of residual plots, multicollinearity, and homoscedasticity.

### Propensity Score Matching and Biomarker-based Classification and Regression Trees

To mitigate selection bias due to incomplete blood sample collection at baseline and to achieve marginal exchangeability between those randomized to the rehabilitation intervention versus attention control who had baseline blood samples for biomarker measurements, we identified matched pairs using the 5 biomarkers followed by demographic covariates and HF subtypes to achieve a 1 to 1 match of participants assigned to rehabilitation intervention or attention control with baseline biomarker measures.^10^ Then we used a classification and regression tree (CART) approach to model within-pair outcome differences to identify easily interpretable subgroup structure.^10, 11^ CART models are extremely flexible and prone to overfit. Trees produced by CART were pruned using Least Squares Means simultaneous confidence intervals to prevent overfitting.^12^ For internal validation of the CART models, given the limited sample size, we used a leave-one-out (LOO) cross validation approach. In this approach, the model is trained on all but one data point, which is withheld for testing. The process is repeated for each data point, leading to multiple model fits that can be used to evaluate model performance and stability.^13^ For more details regarding our regression tree based on propensity score matching approach to model development, we have published the R code.^14^

All tests are two-sided, and the statistically significant level is set at *P*< 0.05. The Inova Health Institutional Review Board reviewed this REHAB-HF sub-study and determined it to be exempt. The REHAB-HF study was approved at each site and written informed consent was obtained including the collection and storage of blood samples for future analysis.

## RESULTS

### Baseline Characteristics According to Treatment Arms

A total of 242 of 349 (69.3%) study participants had baseline biomarkers measured in the trial (**Supplemental Figure 1**). The characteristics of the REHAB-HF participants with and without baseline biomarkers are shown in **Supplemental Table 1**. Participants with baseline biomarkers were more frequently of White race but otherwise had similar demographics and distribution of HF subtypes compared to those participants without biomarkers measured. Demographic, clinical, and biomarker characteristics did not differ significantly between the rehabilitation intervention and attention control treatment groups (**Table 1**). The mean age was 73 years, 52% were female, and 45% were of Black race. At baseline there are no differences between SPPB and 6MWD between the rehabilitation intervention and attention control assigned participants with biomarker measurements. Consistent with advanced age and the acute HF hospitalization, hs-cTnI, hs-cTnT, NT-proBNP, and hs-CRP levels were elevated with mildly reduced eGFR. The correlation between pairs of biomarker concentrations is shown in the heat map in **Supplemental Figure 2.** As expected, there was moderate correlation between all the cardiac specific biomarkers with the greatest correlation between hs-cTnI and T (rho=0.57). Interestingly, hs-CRP wasn’t correlated with any of the more organ specific biomarkers.

**Table 1.** Baseline clinical, biomarker and functional characteristics based on randomized treatment assignment in participants with baseline biomarker measurements. Continuous variables are presented as mean (standard deviation) except biomarkers as median (quartile 1, quartile 3).

| **Clinical Characteristics** | **Attention control**<br>**(n=112)** | **Rehab Intervention**<br>**(n=130)** | **P Value** |
| --- | --- | --- | --- |
| Age (years) | 72 (8) | 73 (8) | >0.9 |
| Female | 64 (57%) | 61 (47%) | 0.12 |
| Black Race | 54 (48%) | 55 (42%) | 0.4 |
| HFpEF | 56 (50%) | 71 (55%) | 0.5 |
| eGFR (mls/min/1.73m<sup>2</sup>) | 56 (23) | 53 (22) | 0.3 |
| **Biomarkers** |  |  |  |
| Hs-cTnT (ng/L) | 34 (23, 49) | 32 (22, 54) | >0.9 |
| Hs-cTnI (ng/L) | 24 (14, 45) | 26 (15, 50) | 0.6 |
| NT-proBNP (ng/L) | 2139 (653, 3929) | 2136 (1089, 4,948) | 0.2 |
| Hs-CRP (mg/L) | 10 (6, 29) | 13(5, 37) | 0.5 |
| Creatinine (mg/dL) | 1.22 (0.96, 1.87) | 1.35 (1.02, 1.72) | 0.4 |
| **Functional Measures** |  |  |  |
| SPPB Score | 6.14 (2.59) | 6.07 (2.74) | 0.8 |
| 6MWD (meters) | 195 (108) | 200 (108) | 0.7 |

Continuous variables are expressed as the mean (standard deviation) except biomarkers as median (quartile 1, quartile 3). 6MWD, 6-minute walk distance; eGFR, estimated glomerular filtration rate; hs-CRP, high-sensitivity C-reactive protein; hs-cTn, high-sensitivity cardiac troponin; HFpEF, heart failure with preserved ejection fraction; NT-proBNP, amino-terminal B-type natriuretic peptide; SPPB, Short Physical Performance Battery.

### Functional Outcomes Predicted by Biomarkers and Interaction with Treatment

Of the 242 participants with baseline biomarker measures, 212 (88%) returned at 12 weeks and had a SPPB measured with 185 (77%) also having a 6MWD test. The mean follow-up SPPB was 8.2±3.1 versus 6.8±3.4 (*p*=0.003) and the mean follow-up 6MWD was 287±127 meters versus 248±136 meters (*p*=0.073) for the rehabilitation intervention and attention control participants, respectively. The number of biomarkers measured at baseline and 12 weeks are shown in the consort diagram in **Supplemental Figure 1**. For SPPB and 6MWD, the association of each biomarker with 12-week change in each functional outcome was evaluated. In **Table 2**, each biomarker was log converted and is shown such that the unadjusted and adjusted point estimates are per log (base 2) change, which represents a doubling of the biomarker level. The consistent trend for both outcomes across all five biomarkers in the overall group of participants (rehabilitation intervention and attention control) is that increasing baseline biomarker levels are associated with less overall subsequent functional improvement by 12-weeks. Several of the baseline biomarker level adjusted associations are statistically significant such that a doubling of baseline NT-proBNP and hs-CRP are associated with a 0.31 (95% confidence intervals [CI] 0.04, 0.58) and 0.47 (95%CI 0.13, 0.80) less improvement in SPPB score respectively controlling for baseline functional outcome measurement. For 12-week change in 6MWD in the adjusted model, NT-proBNP, hs-cTnI and T levels were all associated with less improvement ranging from 14 to 27.2 meters per doubling of the baseline levels. A significant interaction between baseline biomarker level and treatment assignment in the adjusted model is seen for hs-cTnI for the SPPB outcome (*P*=0.04) and hs-cTnT for 6MWD (*P*=0.03). These interactions indicate that baseline biomarker level adjusted associations with SPPB/6MWD are significantly different between the rehabilitation intervention and attention control treatment groups.

**Table 2.** Unadjusted and adjusted linear regression for the association between log (base 2) baseline serum biomarkers with (1) SPPB (Model 1) and (2) 6-minute walk test (Model 2) in all study participants (combined treatment arm) and interactions with randomized treatment arm (Physical Rehabilitation Interventions vs. Attention Control).

|  | Unadjusted*<br>(95% CI) | P-Value | Adjusted**<br>(95% CI) | P-Value | P-Interaction‡ |
| --- | --- | --- | --- | --- | --- |
| **12-week**<br>**change in**<br>**SPPB Score<sup>‡‡</sup>** |  |  |  |  |  |
| Hs-cTnT | -0.23 (-0.58, 0.12) | 0.194 | -0.40 (-0.93, 0.13) | 0.14 | 0.28 |
| **Hs-cTnI** | -0.02 (-0.24, 0.21) | 0.9 | -0.26 (0.58, 0.05) | 0.10 | **0.040** |
| NT-proBNP | -0.09 (-0.29, 0.10) | 0.34 | -0.31 (-0.57, -0.04) | **0.024** | 0.16 |
| Hs-CRP | -0.36 (-0.57, -0.15) | **<0.001** | -0.47 (-0.80, -0.13) | **0.006** | 0.34 |
| Creatinine | -0.48 (-1.12, 0.16) | 0.14 | -0.58 (-1.49, 0.33) | 0.21 | 0.47 |
| **12-week**<br>**change in**<br>**6MWD**<br>**(meters)** |  |  |  |  |  |
| Hs-cTnT | -11.9 (-25.1, 1.2) | 0.08 | -27.2 (-47.4, -7.0) | **0.009** | **0.03** |
| Hs-cTnI | -8.1 (-17.1, 0.8) | 0.07 | -15.5 (-28.5, -2.4) | **0.02** | 0.18 |
| NT-proBNP | -6.7 (-14.2, 0.8) | 0.08 | -14.0 (-24.6, -3.4) | **0.01** | 0.05 |
| Hs-CRP | -6.9 (-15.6, 1.8) | 0.12 | -9.3 (-23.8, 5.1) | 0.20 | 0.62 |
| Creatinine | -20.9 (-46.3, 4.6) | 0.11 | -24.4 (-61.5, 12.8) | 0.20 | 0.51 |

**Abbreviations:** CI = confidence interval; See table 1 for additional abbreviations. \* Estimates are per log2 change in biomarker (essentially per doubling of the biomarker level). †Adjusted for baseline SPPB or 6MWD, intervention type, biomarker level x intervention, age, sex, race category, heart failure category, and intervention x heart failure category. ‡ P-value for interaction product term of baseline biomarker level with intervention type. ‡‡ SPPB scored as a whole number range 1-12. Biomarker outliers removed (hs-cTnT=1, hs-cTnI=2, NT-proBNP=1, hs-CRP=4, creatinine=3).

To visualize changes across baseline biomarker levels and treatment group, **Figures 1 and 2** display the linear associations between log (base 2) baseline biomarker levels and 12-week change in SPPB and 6MWD respectively by treatment assignment. For participants assigned to attention control, the association between all increasing biomarker levels and less functional improvement at 12 weeks is clearly seen for both SPPB and 6MWD. For SPPB, participants assigned to the rehabilitation intervention had consistently better trends for differential improvements in scores at 12-weeks relative to attention control participants with progressively higher baseline NT-proBNP, hs-cTnT, creatinine, and hs-CRP levels. Corresponding to the significant interaction noted above (**Table 2**), as baseline hs-cTnI levels increase, the difference in improvement in scores at 12-weeks between the rehabilitation and attention control participants becomes progressively larger. Similarly, for 6MWD, the rehabilitation intervention participants exhibited consistently better trends for differential improvements in 6MWD at 12-weeks compared to attention control participants with progressively higher levels of baseline creatinine, hs-CRP, hs-cTnI and NT-proBNP; with a significant interaction (**Table 2**) for increasing baseline hs-cTnT and differential improvements in 6MWD at 12-weeks for the rehabilitation intervention participants relative to the attention control participants.

**Figure 1.**
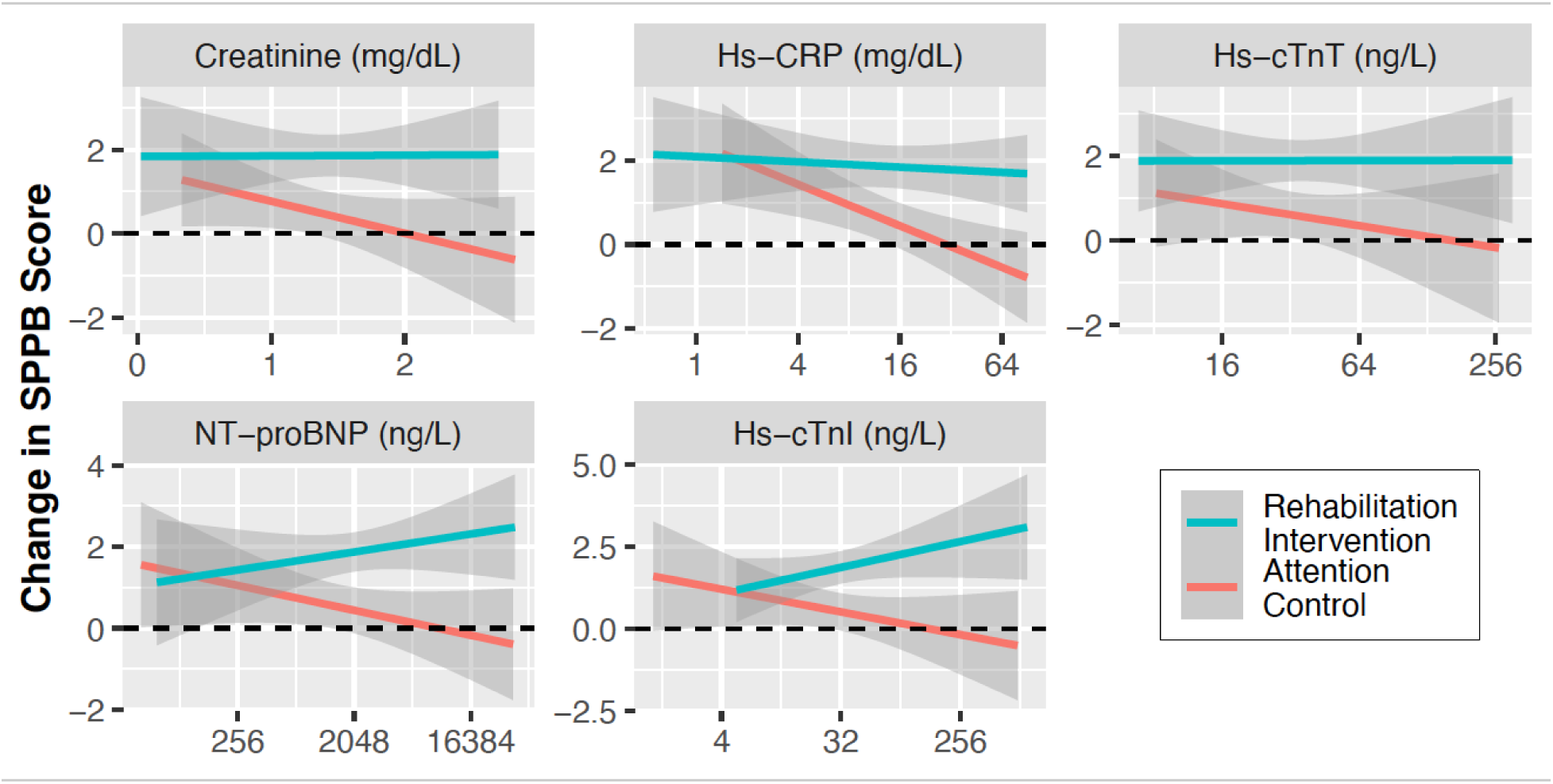
Twelve-week change in Short Physical Performance Battery by Baseline Biomarker Level and Treatment Assignment. Y-axis is the change in Short Physical Performance Battery SPPB from baseline to 12-weeks. The X-axis is the baseline biomarker level shown in concentration units on a log2 scale. Grey areas represent 95% confidence intervals. hs-CRP, high sensitive C-reactive protein; hs-cTnI, high sensitive cardiac troponin I; hs-cTnT, high sensitive cardiac troponin T; NT-proBNP, amino-terminal B-type natriuretic peptide.

**Figure 2.**
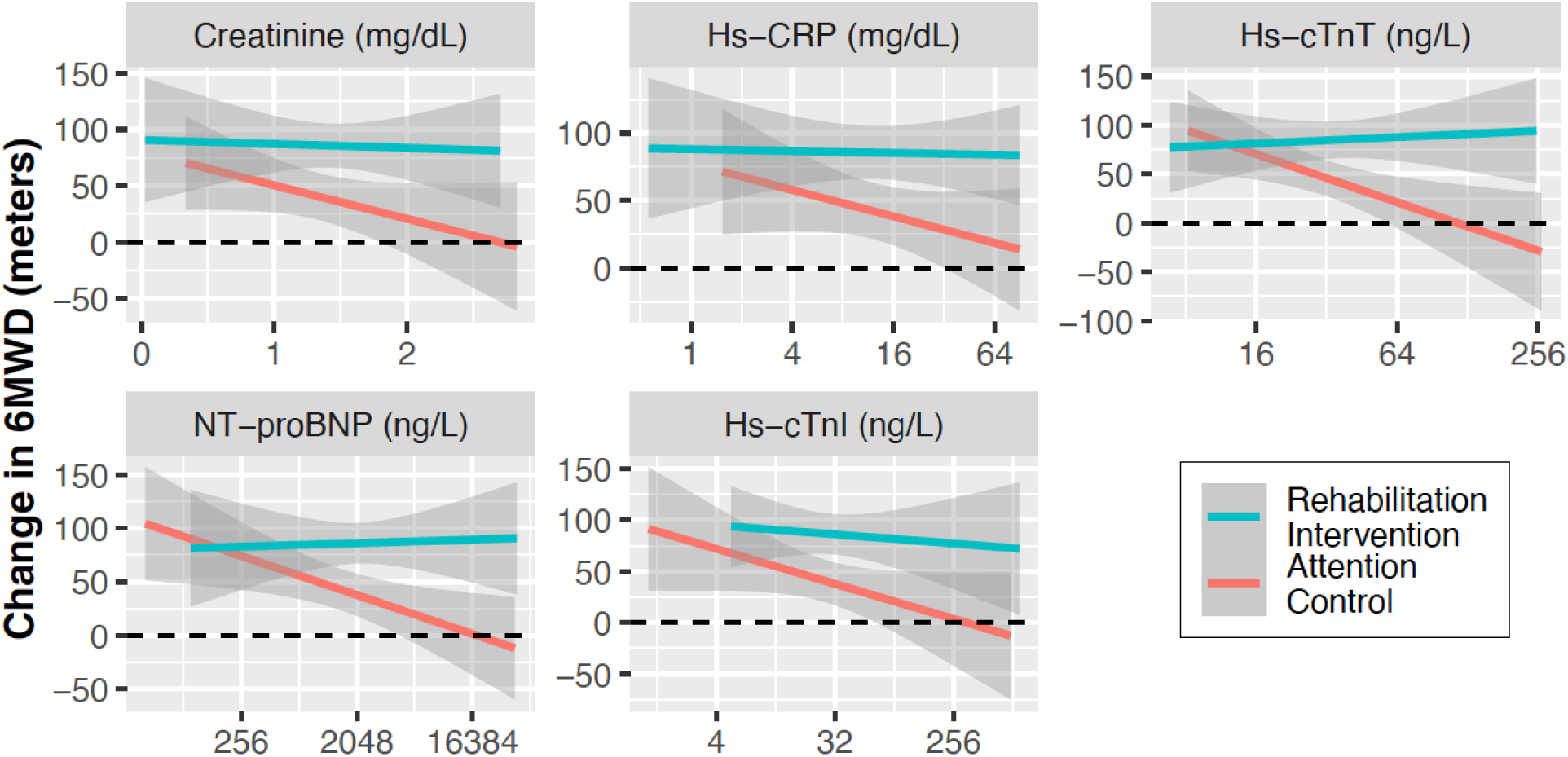
Twelve-week change in Six Minute Walk Distance by Baseline Biomarker level and Treatment Assignment. Y-axis is the change in 6-minute walk distance (6MWD) from baseline to 12-weeks. The X-axis is the baseline biomarker level shown in concentration units on a log2 scale. Grey areas represent 95% confidence intervals. hs-CRP, high sensitive C-reactive protein; hs-cTnI, high sensitive cardiac troponin I; hs-cTnT, high sensitive cardiac troponin T; NT-proBNP, amino-terminal B-type natriuretic peptide.

To explore how baseline biomarker levels could be used together without assuming a constant linear relationship between log (base 2) of the biomarker level and 12-week change in both physical function measures, we performed a CART analysis on propensity score matched participants assigned to either the rehabilitation intervention or attention control. There were 165 participants with both SPPB and 6MWD measured at baseline and 12-weeks who also had all 5 biomarkers measured at baseline (**Supplemental Figure 1**). Seventy-three participants were assigned to attention control and 92 to the rehabilitation intervention. Using the propensity score settings outlined in the methods section, 118 participants were matched (81% of the 146 potential matches [i.e. with 73 attention control participants there could only be 146 matched participants]). The characteristics of the matched (n=118) based on treatment assignment and unmatched (n=47) are shown in **Supplemental Table 2**. There were no differences in the five biomarker levels between matched participants assigned to the rehabilitation intervention versus attention control. Biomarker levels in the unmatched participants were also similar to the matched participants. Demographics and HF subtypes were also not significantly different in the matched pairs assigned to the rehabilitation intervention or attention control. Unmatched participants were also similar to matched participants with respect to demographics and HF subtypes.

Using the propensity score matched pair sample between treatment assignments, we conducted a CART analysis inclusive of the five biomarkers to stratify by differential treatment effect. For SPPB, hs-CRP and creatinine were selected by CART for the tree-based stratification of the treatment effect (**Figure 3a**). Higher hs-CRP (≥9.9 mg/L) and creatinine (≥1.4 mg/dL) differentiated participants assigned to the rehabilitation intervention who had a greater improvement in SPPB at 12-weeks compared to attention control versus participants with lower levels. For optimizing stratification of a differential response based on assignment to the rehabilitation intervention vs attention control for improvement in 6MWD, the CART tree model included hs-CRP with a similar cut-off of ≥9.9 mg/L as SPPB, and further differentiated responders by their hs-cTnT level such that individuals assigned to the rehabilitation intervention with hs-CRP≥9.9 mg/L and a hs-cTnT ≥36 ng/L gained on average 129 (95%CI 78, 180) meters more from baseline to 12-weeks than matched individuals assigned to attention control. In contrast, if the baseline hs-CRP was <9.9 mg/L, the average difference gained was only 20 (95%CI -0.5, 60) meters (**Figure 3b**). The CART 6MWD decision tree model was relatively more stable than the SPPB decision tree model; for the 59 CART models, each constructed by leaving out one of the 59 matched pairs, 55 out of 59 (93%) of 6MWD decision trees had nearly identical tree structures to that of the original model in **Figure 3** built using all of the matched pairs (i.e. a tree structure involving successive splits on hs-CRP and hs-cTnT at values ranging from 9.3-9.9mg/L and 35-36ng/L respectively). Forty one out of 59 (69%) of SPPB decision tree models had nearly identical tree structures to the original model shown in **Figure 3** using all the matched pairs (i.e. involved a tree structure of successive splits on hs-CRP and creatinine at values ranging from 9.4-9.9mg/L and 1.2-1.4 mg/dL respectively).

**Figure 3.**
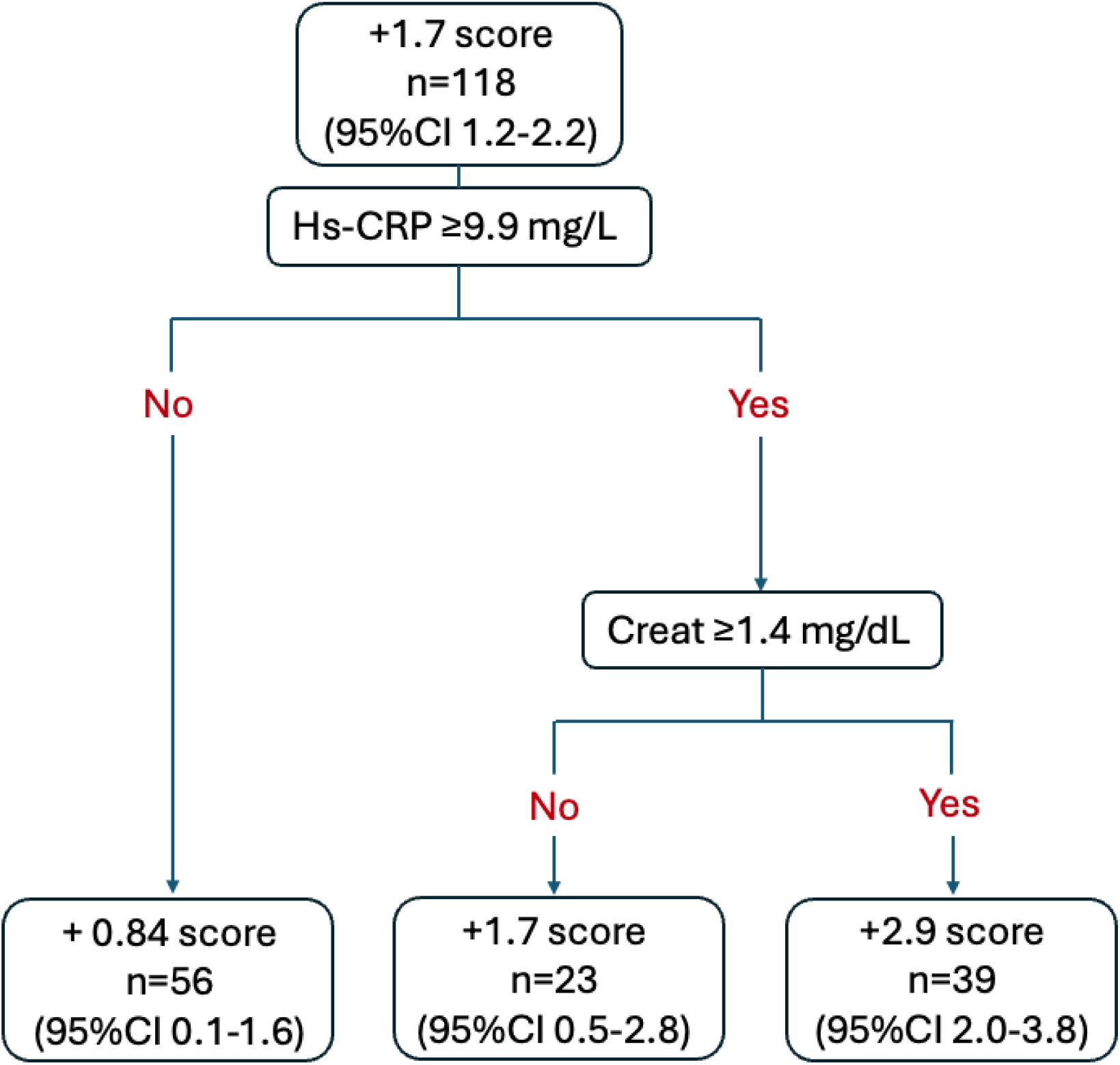
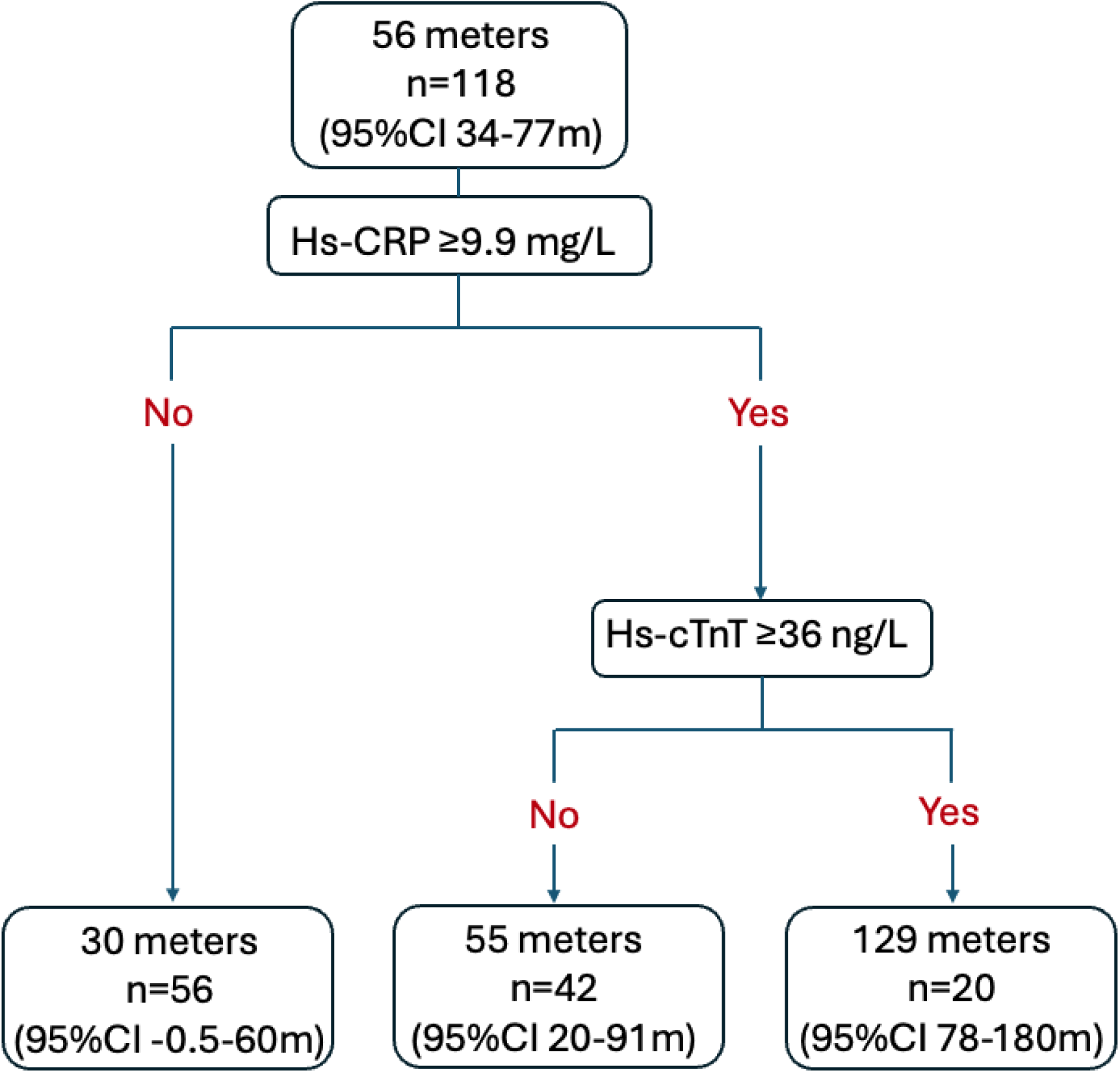
Classification and regression trees utilizing baseline levels of five biomarkers to stratify differential 12-week physical function changes in (**A**) SPPB and (**B**) 6MWD after a heart failure hospitalization based on treatment assignment to either a rehabilitation intervention or attention control. Creat, Creatinine; hs-CRP, high sensitive C-reactive protein; hs-cTnT, high sensitive cardiac troponin T. **A.** Difference in 12-weeks change in Short Physical Performance Battery between the tailored rehabilitation intervention and attention control in propensity score matched participants. **B.** Difference in 12-weeks change in six-minute walk distance between the tailored rehabilitation intervention and attention control in propensity score matched participants.

### Biomarkers and Six-month Clinical Prognosis

We evaluated the baseline levels of the five biomarkers for their association with the 6-month clinical endpoint of major adverse events (all-cause hospitalization and death) and its two components. The number of major adverse events and its components in the 242 participants with baseline biomarkers are shown in **Supplemental Table 3.** There were 28 deaths and 139 all-cause hospitalizations. The composite of the two types of clinical events, or the individual event types did not differ based on treatment assignment. In an adjusted logistic regression analysis, only creatinine was significantly associated with the composite outcome (OR 2.50 [95%CI 1.17-5.52] per doubling of the level), death (OR 7.68 [95%CI 2.15-34.71]) and total rehospitalization (OR 2.28 [95%CI 1.09-5.08]) and had an interaction with treatment and death (*P*=0.02 for interaction). NT-proBNP was associated with death with an adjusted OR 1.67 (95%CI 1.14-2.61). The unadjusted and adjusted odds ratios for each biomarker and each outcome as well as the interaction with the biomarkers and treatment for the outcome are shown in **Supplemental Table 4**.

### Change in Biomarker Levels from Baseline to 12-weeks

The change in each of the five biomarkers stratified based on treatment assignment to the rehabilitation intervention versus attention control is shown in the violin plots in **Supplemental Figure 3**. All biomarkers except creatinine significantly decreased from baseline to 12-weeks The median (quartile 1, quartile 3) change from baseline to 12-weeks for the biomarkers was as follows: hs-cTnT -0.9 (5.5, - 9.7) ng/L (*P*=0.002), hs-cTnI -3.2 (4.5, -16.8) ng/L (*P*<0.001), NT-proBNP -162 (453, -1192) ng/L (*P*=0.012), hs-CRP -4.3 (-0.3, -21.3) mg/L (*P*<0.001), creatinine 0.04 (0.23, -0.12) mg/dL (*P*=0.61). However, no biomarker level differentially changed more based on treatment assignment.

## DISCUSSION

In this secondary analysis of the REHAB-HF study, which included biomarkers from various physiological domains, we report several key findings: (1) near the time of discharge for acute HF hospitalization, many participants had elevated biomarkers indicating cardiac injury, congestion, general inflammation, and impaired renal function. These elevations reflect the accelerated “pathological aging” and severe acute cardiovascular decompensation in this older, often frail population; (2) the multidomain early tailored progressive rehabilitation intervention showed stable efficacy across progressively higher biomarker levels for both the SPPB and the 6MWD. This suggests that despite the biochemical evidence of decompensated cardiovascular disease, the rehabilitation intervention remained effective and applicable; (3) in contrast to the rehabilitation intervention, participants assigned to the attention control with higher baseline biomarker levels showed little to no improvement in functional outcomes; (4) the CART analysis showed that specific biomarkers could be used together to optimize the stratification of treatment efficacy by utilizing personalized treatment approaches based on biomarker profiles; and (5) the biomarkers did not selectively change over 12-weeks based on treatment assignment. This indicates that the effectiveness of the rehabilitation intervention in improving physical function is not due to selective reductions in cardiac injury, congestion from heart failure, improvements in renal function, or reductions in generalized inflammation.

Older adults often live with age-associated conditions called “geriatric syndromes”. Geriatric syndromes are a group of clinical conditions that do not fit into a discrete disease categories.^15^ These syndromes are frequently the result of accelerated “pathologic” aging due to chronic morbidity, inflammation, metabolic derangement, coagulopathy, and other biological mechanisms that result in worse health outcomes, particularly following acute cardiovascular illness.^15^ These syndromes include frailty, frequent falls, delirium, cognitive impairment, multimorbidity, polypharmacy, and functional decline.^15^ Older cardiovascular disease patients, including those with HF, are at the greatest risk for developing geriatric syndromes, and a multidisciplinary approach to geriatric conditions that includes early screening and tailored physical rehabilitation intervention significantly improve physical function, frailty status, and quality of life in older adults with HF.^4^ The REHAB-HF trial showed that pre-frail and frail patients admitted with ADHF had improvement in SPPB, 6MWT, gait speed, frailty status, and overall quality of life at 3-months.^4^ The effect of the multidomain rehabilitation intervention can extend cardiovascular management to address geriatric conditions including physical decline and frailty, particularly after acute cardiovascular illness.

Biomarkers play a role in identifying physiologic mechanisms that can be explained by acute cardiovascular illness and pathological aging. We observed that biomarkers were elevated near discharge with an acute HF admission, and importantly higher levels of these biomarkers are associated with less functional improvement by 12 weeks when treated with attention control. This study also reveals the nuanced interaction between treatment assignment and biomarker levels. Specifically, the rehabilitation intervention group showed consistent efficacy across progressive elevation of all baseline biomarker levels with respect to improvement in SPPB and 6MWT compared to participants randomized to attention control where functional improvement declined with progressively higher baseline biomarker levels. This suggests that the early, tailored and progressive multidomain physical rehabilitation intervention may mitigate many of the adverse effects associated with elevated biomarker levels representing multiple pathophysiologic domains, thereby enhancing recovery in particularly vulnerable older adults.

The CART analysis provides an approach that improves clinical interpretation and further supports the utility of multiple baseline biomarkers in stratifying treatment effects. Participants with higher levels of hs-CRP and creatinine at baseline demonstrated greater improvements in SPPB scores when assigned to the rehabilitation intervention, compared to attention control. In pre-clinical models inflammation is a consistent factor in the progression to frailty.^16^ In a prespecified secondary analysis of REHAB-HF where participants were dichotomized as pre-frail or frail using the Fried criteria there was a significant interaction with the presence of frailty and SPPB such that frail, versus pre-frail participants assigned to the rehabilitation intervention had a greater differential improvement in SPPB compared to attention control.^17^ Therefore, the continuous measure of differentiation of improvement in SPPB based on hs-CRP levels provides consistency with the clinical assessment of frailty. Similarly, for 6MWD, participants with higher hs-CRP and hs-cTnT levels showed greater differential improvement from the rehabilitation intervention. Hs-cTnT can be used to predict incident HF in older adults, and predicts prognosis following acute HF admission.^18, 19^ Measures of renal function and hs-CRP are also well understood to be prognostic in patients with prevalent HF.^20, 21^ We extend these findings to underscore the potential for these biomarkers to guide clinical decision-making and optimize treatment strategies, particularly with a broadly pleotropic treatment such as a multidomain tailored physical therapy intervention in geriatric populations where the interplay of multiple health factors can complicate management.

The biomarker hs-cTnT has been associated with both cardiac and skeletal muscle damage.^22^ Prior work has identified that higher circulating hs-cTnT elevations were found in patients with underlying noninflammatory skeletal muscle myopathy and myositis.^23^ The hypothesis was that damaged skeletal muscle was implicated in higher systemic concentration of hs-cTnT, but not hs-cTnI, a biomarker that can be more specific to cardiac muscle cells in the presence of skeletal muscle dysfunction.^22^ cTnT expression is also measurable in the skeletal muscle of older adults without overt myopathies.^24^ Older patients with cardiovascular conditions have a higher prevalence of skeletal muscle mass disorder than younger patients.^25^ Skeletal muscle disorders are often exacerbated by the presence of multimorbidity, drug-drug or drug-disease interactions, chronic inflammation, oxidative stress, insulin resistance, and endothelial dysfunction.^26, 27^ Specifically, patients with HFpEF have a greater extent of skeletal muscle mitochondrial dysfunction compared to age matched older adult controls.^28^ Thus, elevated levels of hs-cTnT may reflect underlying HF and age associated risks including geriatric syndromes and offers a physiologic explanation for the statistical interaction with treatment assignment and 6MWD, a metric that reflects both cardiovascular and skeletal muscle fitness.

This study provides additional evidence that baseline biomarkers are significant predictors of functional outcomes and clinical prognosis in older adults with recent acute HF hospitalizations. While the traditional approach to measuring outcomes in cardiovascular trials were focused on major adverse cardiovascular events (MACE) outcomes, a shift towards a more patient-centered approach to include functional outcomes is necessary for older adults.^29^ The differential improvements observed with the rehabilitation intervention compared with attention control underscore the importance of targeting those with highest biomarker levels with the rehabilitation intervention because they derive the most benefit from the intervention addressing the unique needs of geriatric patients. This is particularly important for clinicians to recognize that these patients potentially with the greatest comorbidities with engagement in a 12-week outpatient multidomain rehabilitation intervention program stand to derive the greatest relative benefit. Future research should continue to explore the integration of biomarker assessments in clinical practice, aiming to refine treatment algorithms and improve outcomes for older adults with HF. A holistic and multidisciplinary approach remains essential in managing the complexities of geriatric syndromes and optimizing care for this growing population.

### Study Strengths and Limitations

The REHAB-HF trial is unique because it enrolled older adults admitted with acute HF and randomized to an early, transitional, tailored, progressive multidomain rehabilitation intervention that improved physical function, frailty, and quality of life metrics. With the advancing age of the U.S. population, clinical overt and subclinical skeletal muscle disorders are increasingly prevalent in cardiovascular practice.^25^ This study measured circulating biomarkers in the context of acute rehabilitation intervention at baseline and follow-up and addressed important issues related to the biomarkers as effect measure modifiers of treatment on domains of physical function. The use of these clinically available biomarkers offers analytically precise continuous measures versus less precise categorical clinical assessment for physiologic insights into these post-acute HF participant response to the rehabilitation intervention versus attention control. The study has important limitations. The presence of a significant interaction with treatment or selection in the CART analysis does not imply biologic causality for a role of the efficacy of the rehabilitation intervention. In fact, we saw no differential change at 12-weeks in any of the biomarkers as a result of intervention assignment. However, given that all the biomarkers except creatine decreased over 12-weeks, it is possible that the effect of ongoing convalescence and decongestion following the acute HF hospitalization may have overwhelmed a causal biologic change resulting from the physical therapy intervention. Future studies enrolling older patients in rehabilitation intervention should consider the utility of studying skeletal muscle directly through imaging and biopsies which may provide more mechanistic findings to support the statistical associations seen with the biomarkers and physical function outcomes in REHAB-HF. Second, selection bias is potentially present in those who had biomarkers measured in this study. This is noted demographically by differences in distribution of race in those with and without biomarkers measured and could impact the exchangeability obtained from randomization. We addressed this issue by propensity score matching participants with baseline biomarker measures to re-establish exchangeability between treatment assignments, but we cannot exclude an imbalance due to unmeasured confounders. The use of CART has the advantage that we didn’t need to assume a linear relationship between the biomarkers and outcomes. We further attempted to validate our CART models using a leave one out cross validation approach to assess stability and performance. With 69% of the model replications similar to the primary SPPB tree model, this suggests the possibility that other partitions of the data identifying subgroups with heterogeneous treatment effects may also exist. However, given the limited sample size, moderate correlation among baseline biomarkers, and the nature of CART’s greedy search algorithm for building tree-based models^11^, it is not surprising that the exact tree structure cannot be recovered in all leave-one-out models.^30, 31^ The generalizability of our findings is also limited until they can be externally validated in a different cohort. This will be possible after the completions of the REHAB-HFpEF study (Clinical Trials.Gov ID: NCT05525663). The REHAB-HF study population includes only older adults with acute heart failure. The results of this study may not be generalized to other forms of acute CVD including acute coronary syndrome and valvular heart disease, or those with symptomatic prevalent CVD.

## CONCLUSION

This secondary analysis of the REHAB-HF, a study of older adults recently hospitalized with acute heart failure assigned to a multidomain physical rehabilitation intervention versus attention control, found elevated biomarkers at discharge, indicative of cardiac injury, congestion, inflammation, and renal impairment, consistent with pathological aging and cardiovascular decompensation prevalent in this frail population. Despite high levels of these biomarkers, the multidomain rehabilitation intervention consistently demonstrated efficacy across varying degrees of disease severity, as reflected in stable improvements in the SPPB and 6MWD. Our CART analysis reveals the potential for using specific biomarkers to optimize treatment stratification, thereby enhancing the personalization and effectiveness of rehabilitation interventions.

## Data Availability

"All data referred to in this manuscript are subject to restrictions imposed by the funders and will not be publicly available. However, all R output and associated code used for the analysis have been made publicly accessible to ensure transparency and reproducibility.

https://github.com/sbruce23/RehabHFbiomarkers

---

**Supplemental Table 1.** Clinical characteristics of REHAB-HF participants with and without biomarkers measured at baseline. Continuous variables are presented as mean (standard deviation).

| **Clinical Characteristics** | **Baseline Biomarkers measured (n=242)** | **No baseline biomarkers measured (n=107)** | **P Value** |
| --- | --- | --- | --- |
| Age (years) | 72(8) | 73(8) | 0.6 |
| Female | 125(52%) | 58(54%) | 0.7 |
| Non-White Race | 109(45%) | 63(59%) | 0.02 |
| HFpEF | 127(52%) | 58(54%) | 0.8 |
| eGFR (mls/min/1.73m<sup>2</sup>) | 54(22) | 55(22) | 0.6 |
| **Functional Measures** |  |  |  |
| SPPB Score | 6.10(2.67) | 5.98(2.77) | 0.8 |
| 6MWD (meters) | 198(108) | 183(99) | 0.3 |
| 6MWD, 6-minute walk distance; eGFR, estimated glomerular filtration rate; SPPB, Short Physical Performance Battery. |  |  |  |

**Supplemental Table 2.** Baseline clinical and biomarker characteristics based on propensity score selected matched pair participants and unmatched participants who had all five biomarkers measured at baseline and both an SPPB and 6MWD measured at baseline and 12-weeks.

|  | **Propensity score matched** |  | **Unmatched** |
| --- | --- | --- | --- |
| **Clinical Characteristics** | Attention control (n=59) | Rehab Intervention (n=59) | n=47 |
| Age (years) | 71(8) | 72(8) | 72(7) |
| Female | 33(56%) | 24(41%) | 25(53%) |
| Non-White Race | 24(41%) | 20(34%) | 27(57%) |
| HFpEF | 24(41%) | 29(49%) | 20(43%) |
| eGFR<br>(mls/min/1.73m<sup>2</sup>) | 59(22) | 55(21) | 55(26) |
| **Biomarkers** |  |  |  |
| Hs-cTnT (ng/L) | 33 (23, 44) | 32 (22, 42) | 29 (19, 72) |
| Hs-cTnI (ng/L) | 24 (16, 42) | 25 (17, 40) | 28 (12, 111) |
| NT-proBNP (ng/L) | 1,937 (760, 3,480) | 1,892 (1,056, 4,303) | 1,937 (805, 4,502) |
| Hs-CRP (mg/L) | 10 (6, 26) | 12 (5, 27) | 11 (4, 41) |
| Creatinine (mg/dL) | 1.19 (0.97, 1.44) | 1.29 (1.02, 1.62) | 1.32 (0.99, 1.90) |
| Continuous variables are expressed as the mean (standard deviation) except biomarkers as median (quartile 1, quartile 3). 6MWD, 6-minute walk distance; eGFR, estimated glomerular filtration rate; hs-CRP, high-sensitivity C-reactive protein; hs-cTn, high-sensitivity cardiac troponin; HFpEF, heart failure with preserved ejection fraction; NT-proBNP, amino-terminal B-type natriuretic peptide; SPPB, Short Physical Performance Battery. |  |  |  |

**Supplemental Table 3.** Clinical six-month outcomes by treatment assignment in the REHAB-HF Trial among participants who had biomarkers measured at baseline.

| **Variable** | **N** | **Attention control**<br>**(n=112)** | **Rehab Intervention**<br>**(n=130)** | ***P*-Value** |
| --- | --- | --- | --- | --- |
| Rehospitalization or Death | 242 | 69 / 112 (62%) | 75 / 130 (58%) | 0.6 |
| Rehospitalization | 242 | 67 / 112 (60%) | 72 / 130 (55%) | 0.5 |
| Death | 242 | 12 / 112 (11%) | 16 / 130 (12%) | 0.8 |
| Lost to follow up | 242 | 7 / 112 (6.3%) | 9 / 130 (6.9%) | >0.9 |

**Supplemental Table 4.**
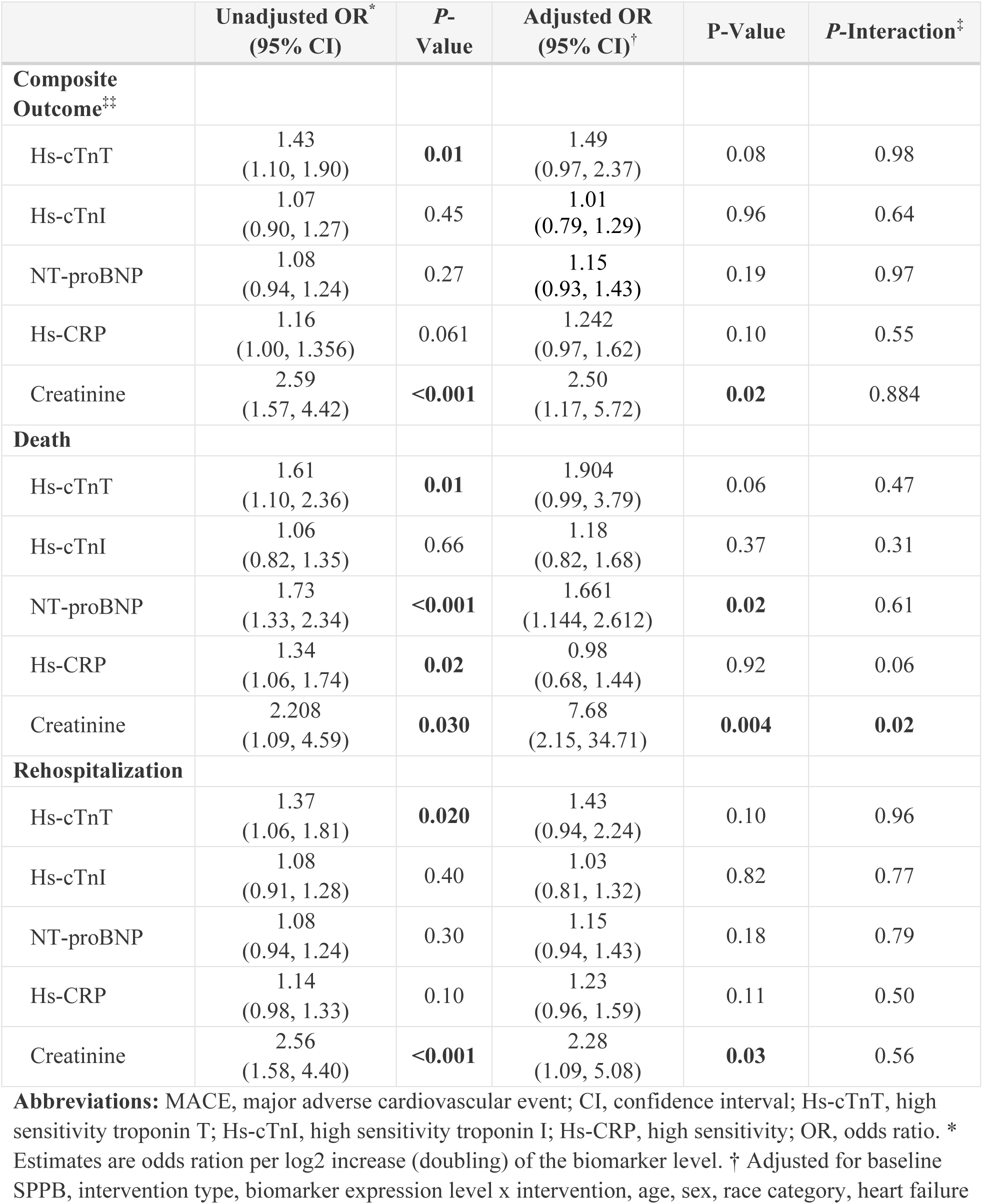
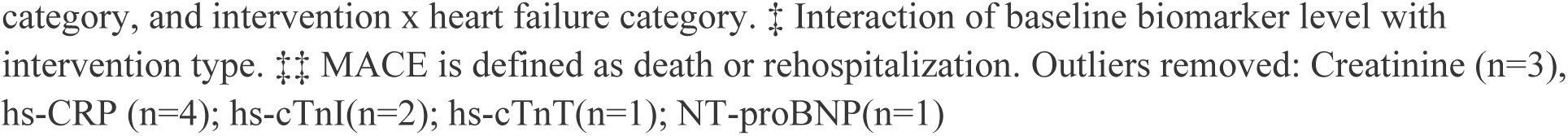
Unadjusted and adjusted logistic regression for association between baseline serum biomarkers with six-month clinical outcomes.

|  | **Unadjusted OR\***<br>**(95% CI)** | **P-**<br>**Value** | **Adjusted OR**<br>**(95% CI)†** | **P-Value** | **P-Interaction‡** |
| --- | --- | --- | --- | --- | --- |
| **Composite Outcome<sup>‡‡</sup>** |  |  |  |  |  |
| Hs-cTnT | 1.43<br>(1.10, 1.90) | **0.01** | 1.49<br>(0.97, 2.37) | 0.08 | 0.98 |
| Hs-cTnI | 1.07<br>(0.90, 1.27) | 0.45 | 1.01<br>(0.79, 1.29) | 0.96 | 0.64 |
| NT-proBNP | 1.08<br>(0.94, 1.24) | 0.27 | 1.15<br>(0.93, 1.43) | 0.19 | 0.97 |
| Hs-CRP | 1.16<br>(1.00, 1.356) | 0.061 | 1.242<br>(0.97, 1.62) | 0.10 | 0.55 |
| Creatinine | 2.59<br>(1.57, 4.42) | **<0.001** | 2.50<br>(1.17, 5.72) | **0.02** | 0.884 |
| **Death** |  |  |  |  |  |
| Hs-cTnT | 1.61<br>(1.10, 2.36) | **0.01** | 1.904<br>(0.99, 3.79) | 0.06 | 0.47 |
| Hs-cTnI | 1.06<br>(0.82, 1.35) | 0.66 | 1.18<br>(0.82, 1.68) | 0.37 | 0.31 |
| NT-proBNP | 1.73<br>(1.33, 2.34) | **<0.001** | 1.661<br>(1.144, 2.612) | **0.02** | 0.61 |
| Hs-CRP | 1.34<br>(1.06, 1.74) | **0.02** | 0.98<br>(0.68, 1.44) | 0.92 | 0.06 |
| Creatinine | 2.208<br>(1.09, 4.59) | **0.030** | 7.68<br>(2.15, 34.71) | **0.004** | **0.02** |
| **Rehospitalization** |  |  |  |  |  |
| Hs-cTnT | 1.37<br>(1.06, 1.81) | **0.020** | 1.43<br>(0.94, 2.24) | 0.10 | 0.96 |
| Hs-cTnI | 1.08<br>(0.91, 1.28) | 0.40 | 1.03<br>(0.81, 1.32) | 0.82 | 0.77 |
| NT-proBNP | 1.08<br>(0.94, 1.24) | 0.30 | 1.15<br>(0.94, 1.43) | 0.18 | 0.79 |
| Hs-CRP | 1.14<br>(0.98, 1.33) | 0.10 | 1.23<br>(0.96, 1.59) | 0.11 | 0.50 |
| Creatinine | 2.56<br>(1.58, 4.40) | **<0.001** | 2.28<br>(1.09, 5.08) | **0.03** | 0.56 |

**Abbreviations:** MACE, major adverse cardiovascular event; CI, confidence interval; Hs-cTnT, high sensitivity troponin T; Hs-cTnI, high sensitivity troponin I; Hs-CRP, high sensitivity; OR, odds ratio. \* Estimates are odds ratio per log2 increase (doubling) of the biomarker level. † Adjusted for baseline SPPB, intervention type, biomarker expression level x intervention, age, sex, race category, heart failure
category, and intervention x heart failure category. ‡ Interaction of baseline biomarker level with intervention type. ‡‡ MACE is defined as death or rehospitalization. Outliers removed: Creatinine (n=3), hs-CRP (n=4); hs-cTnI(n=2); hs-cTnT(n=1); NT-proBNP(n=1)

**Supplemental Figure 1.**
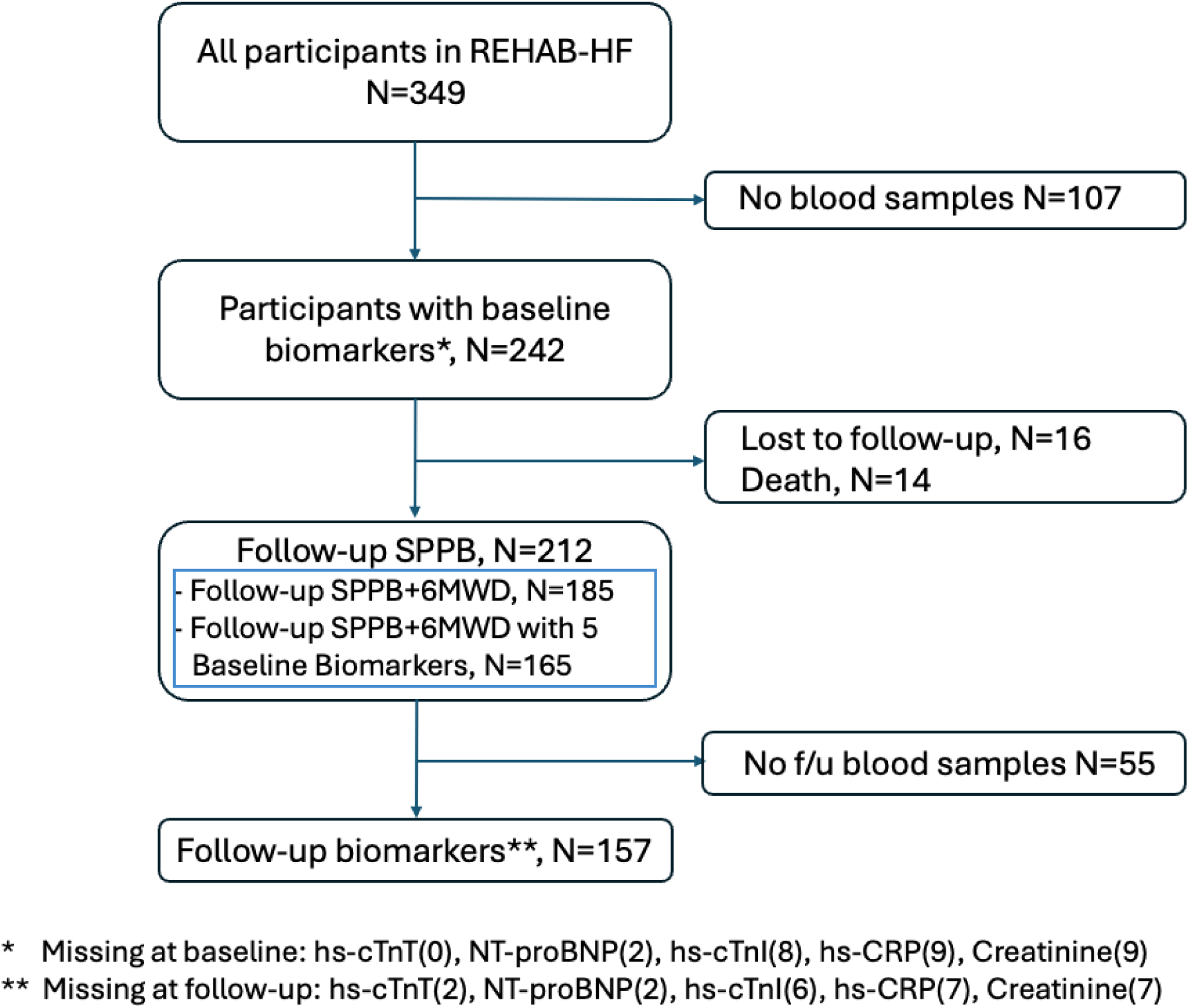
Consort diagram of REHAB-HF participants with baseline and follow-up measures of five circulating biomarkers.

**Supplemental Figure 2.**
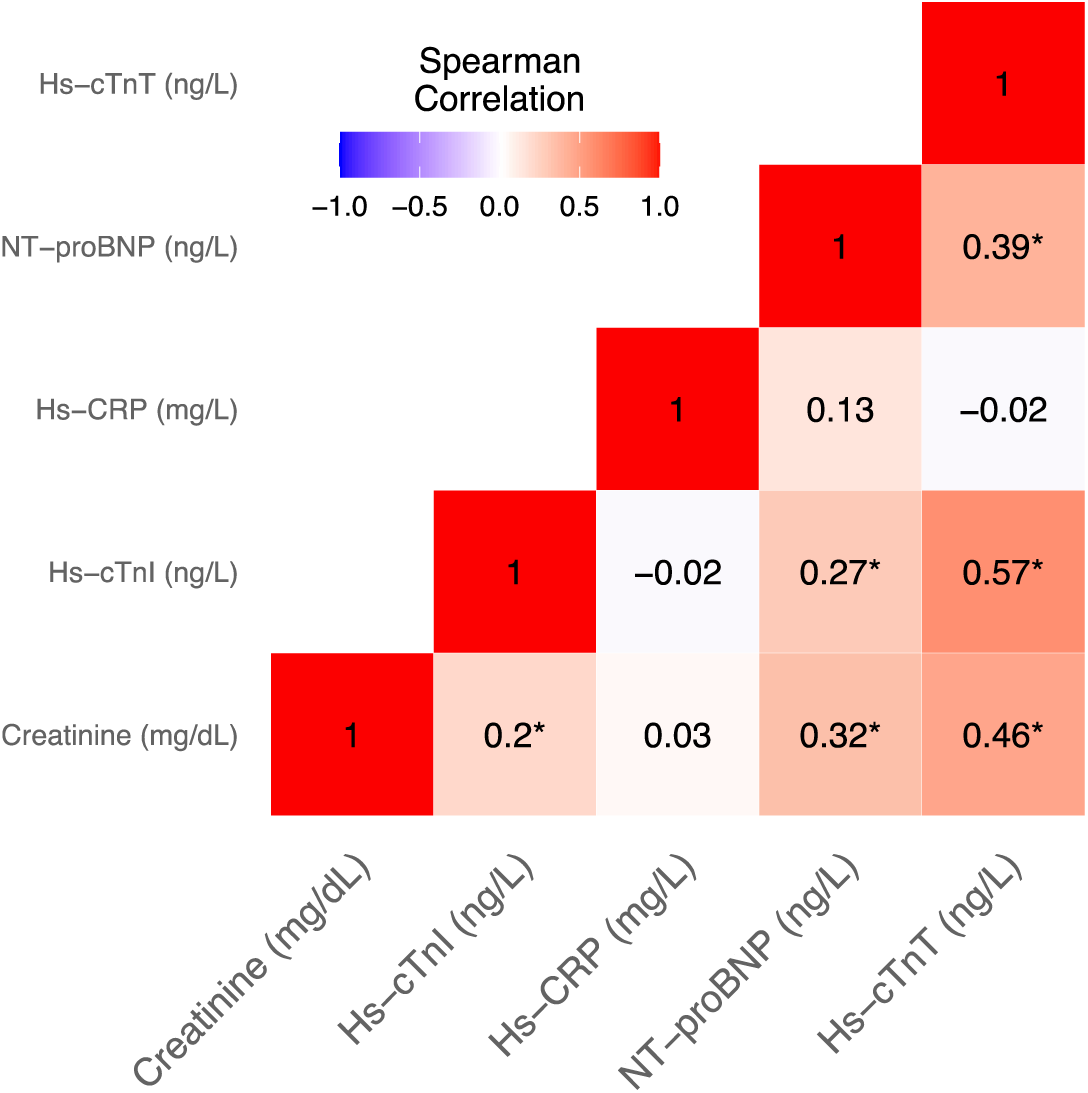
Spearman correlation matrix in pairs of biomarker concentrations.

**Supplemental Figure 3.**
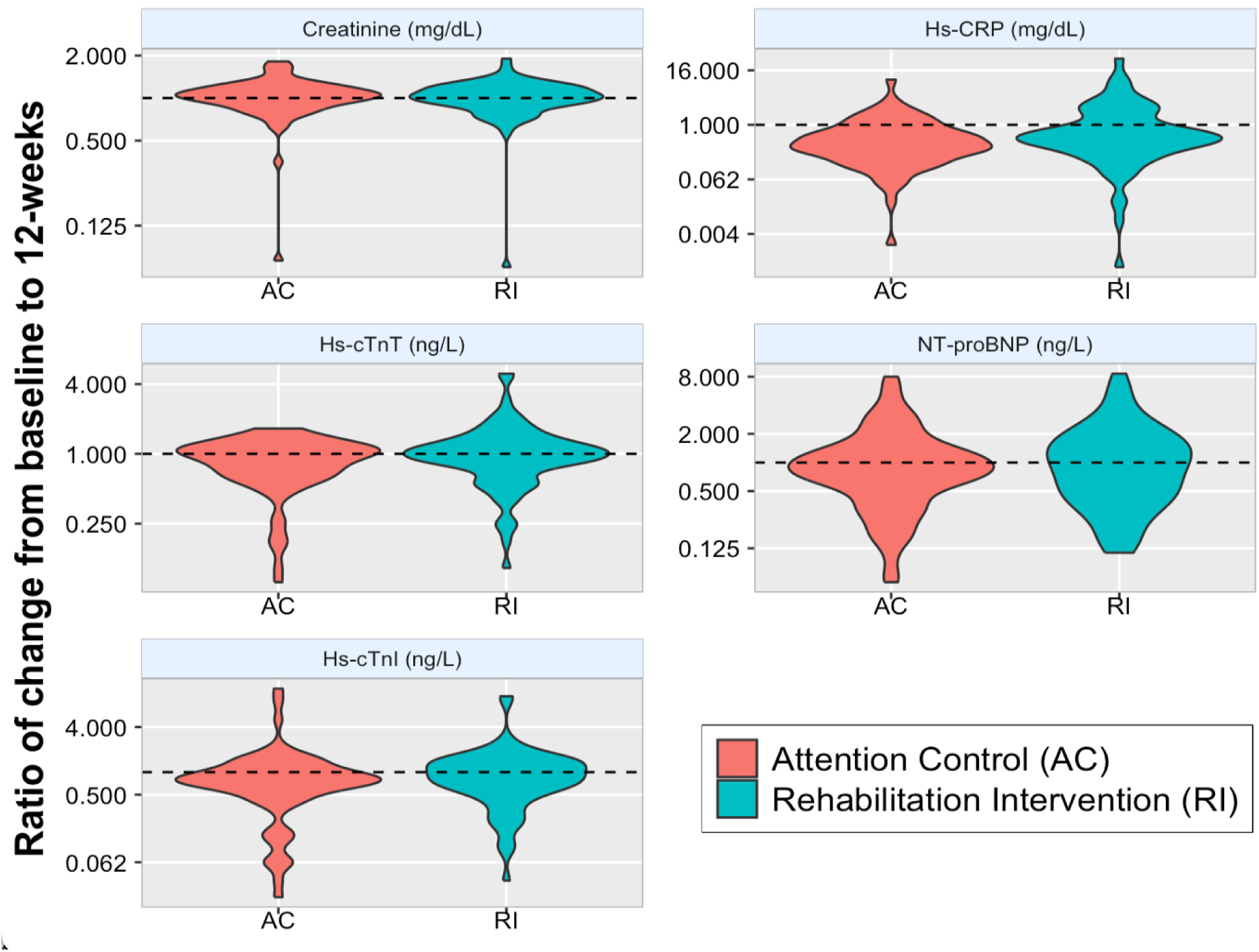
Violin plots of change from baseline to 12-weeks post hospitalization for the five measured circulating biomarkers stratified by treatment assignment to the rehabilitation intervention or attention control.

